# Social patterning of elevated blood lead among children aged 12–59 months in Bangladesh: a national cross-sectional analysis of MICS7 2025

**DOI:** 10.64898/2026.10.09.26365102

**Authors:** Syed Toushik Hossain

## Abstract

**Background:** Lead has no safe level in children. The 2025 Bangladesh Multiple Indicator Cluster Survey (MICS7) provides the first nationally representative blood lead data for the country. Using the WHO Commission on Social Determinants of Health framework, we examined the social patterning of elevated blood lead (≥ 5 µg/dL).

**Methods:** Of 18,905 age-eligible children, 10,622 had a valid blood lead measurement and 9,717 complete covariates. We fitted survey-weighted logistic (aOR) and modified Poisson (aPR) models with 20 covariates across six domains. Biomarker non-response (43.8%), as expected for pediatric venous sampling, was addressed through inverse-probability-of-selection weighting, multiple imputation (*m* = 20), selection-bias scenarios and 500 Rao–Wu bootstrap replicates; the IPW-adjusted prevalence exceeded the naive estimate, suggesting the naive estimate may be conservative under the specified selection model, although this direction is model-dependent.

**Results:** Weighted prevalence was 38.5% (95% CI 37.2–39.9) at ≥ 5 µg/dL and 66.2% at the CDC reference value (≥3.5 µg/dL). Prevalence rose from 30.2% in the poorest to 51.9% in the richest wealth quintile (aOR 1.24 per quintile; 1.13–1.37). Maternal education (aOR 0.83 per level; 0.79–0.88) and ownership of agricultural land (0.77; 0.67–0.88), livestock (0.77; 0.67–0.88) and the dwelling (0.54; 0.43–0.69) were inversely associated. The wealth association differed across divisions (Wald 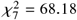, *p* < 0.001), strongest in Dhaka (aOR 1.70; 1.52–1.90). The wealth × maternal-education product term was significant (aOR 0.945; 0.913–0.977), and the wealth association remained positive at higher maternal education (per-quintile aPR 1.09; 1.02–1.16 at higher secondary). The wealth association was robust to most selection-bias scenarios, but attenuated under two adverse asymmetric scenarios and crossed the null under a symmetric worst case (aOR 0.915; 0.833–1.006).

**Conclusions:** Elevated blood lead follows a reverse socioeconomic gradient: wealthier children are more exposed, while maternal education and agrarian assets are inversely associated. This is a social pattern, not a source attribution; market-mediated pathways remain hypotheses. Surveillance should extend beyond poverty-targeted frameworks while continuing to protect the poorest.

## 1 Background

Lead exposure is a well-established global public health concern due to its high toxic effects on the human body [1–3]. Elevated exposure to lead reduces IQ and cognitive development in children [4–6], and is associated with anemia [1]; in adults it is also associated with kidney damage [7] and cardiovascular disease [8]. Lead poisoning is more severe in the pediatric population, as children are more vulnerable and their protective health barriers are not fully developed. Absorption of ingested lead is substantially higher in young children than in adults; infant balance studies reported retention of roughly 40% of an ingested dose, several-fold the fraction usually assumed for adults [9], although the magnitude of this difference in older children is less certain [10]. The World Health Organization lists lead among ten chemicals of major public health concern [1], and no blood lead level in children has been identified as safe [11,12]. Children from low- and middle-income countries (LMICs) bear a disproportionate share of the global burden of lead exposure [13–15], a pattern commonly attributed to weaker regulatory enforcement, environmental monitoring and clinical surveillance.

Past decades of research conducted in high-income countries have yielded a persistent epidemiological result of lead exposure: marginalized and low-income households are more exposed to lead due to conditions such as poor housing and environmental exposures (e.g., lead in air, soil, and water) [16–19]. This inverse socioeconomic gradient has historically underpinned surveillance priorities and mitigation programs focused on poorer households [12, 17].

Whether this inverse gradient among marginalized populations fits in rapidly industrializing economies remains an open and consequential question. Exposure pathways documented in LMICs include adulterated spices [20–22], consumer products such as cookware, toys, paints and cosmetics [23], and informal used lead-acid battery recycling [13, 24]. Critically, several market-based pathways may intensify as household consumption rises, which could produce a positive association rather than an inverse socioeconomic gradient in some contexts. Whether affluent households are in fact more exposed to market-based sources has not, to our knowledge, been tested at national scale; we therefore treat a positive wealth gradient as a hypothesis that the MICS7 data can examine.

We therefore analyzed that hypothesis in the Bangladesh-specific context. Bangladesh faces one of the world’s highest burdens of childhood lead toxicity [15, 25]. Prior Bangladeshi studies documented elevated blood lead levels (BLLs) among Dhaka schoolchildren (mean 15.0 µg/dL) [26], near the Tongi industrial belt (99% above 10 µg/dL) [27], in Dhaka and rural Dinajpur (54% above 10 µg/dL) [28], and around a former lead-acid battery recycling site in Savar, where distance to the site was the strongest predictor [24]. Yet these studies remain location-specific, and their convenience sampling frames make it difficult to establish national determinants.

The 2025 Bangladesh Multiple Indicator Cluster Survey (MICS7) now provides the first nationally representative blood lead measurements in children aged 12–59 months [29, 30]. Because venous biomarker collection in young children is logistically challenging and the blood lead module is a recent addition to the survey instrument, a substantial proportion of age-eligible children did not provide a valid sample, motivating the prespecified non-response framework described below. We analysed these data with four objectives: (1) to characterise the distribution and prevalence of elevated blood lead across structural and intermediate social determinants, using the WHO Commission on Social Determinants of Health (CSDH) framework; to test whether household wealth follows the expected inverse relationship with child lead burden or inverts; (3) to examine whether maternal education and household wealth jointly shape exposure; and (4) to quantify sub-national geographic variation, including heterogeneity of the wealth association, and to assess the sensitivity of findings to non-response and selection. The official MICS 2025 reports present descriptive tabulations of prevalence by background characteristics [29, 30]; our contribution is to apply a formal CSDH framework, provide the first formal test of divisional heterogeneity of the wealth–BLL association using nationally representative data, and characterise wealth ×maternal-education interaction on both multiplicative and additive scales.

## 2 Methods

### 2.1 Data source

We conducted a secondary analysis of the 2025 Bangladesh MICS7, implemented by the Bangladesh Bureau of Statistics with technical assistance from UNICEF [29]. MICS7 used a two-stage stratified cluster sampling design based on the national population census. Venous blood was drawn from children aged 12–59 months and transferred to a designated national laboratory for quantification of blood lead; the Institute of Epidemiology, Disease Control and Research (IEDCR) and the International Centre for Diarrhoeal Disease Research, Bangladesh (icddr,b) were technical partners for specimen collection, cold-chain transport and laboratory testing [29, 30]. Field-level quality assurance included trained phlebotomists, direct supervision and CAPI-based data entry [29]. All analyses incorporated primary sampling units (PSUs), strata and child blood lead sample weights (chblweight). The study is reported according to the STROBE statement [31]; the checklist is Additional file 3.

The final MICS7 Survey Findings Report reports 10,661 children aged 12–59 months with a completed blood lead measurement and a national prevalence of 38.4% at ≥5 µg/dL [29]. Our analytic sample contains 10,622 children (99.6% of the report’s total); the 39-child difference reflects children whose laboratory result was rejected or whose specimen barcode did not match, whom we treat as missing rather than as not-elevated. Under the report’s denominator convention, our national estimate would be 38.4% rather than 38.5%, and the Dhaka division estimate would be 65.2% rather than 65.6%, differences well within sampling error. All national and division-level descriptive estimates agree with the report to within 0.4 percentage points (Additional file 1, Table S3).

### 2.2 Study population and non-response

The analytic population comprised children aged 12– 59 months with a valid blood lead measurement; the derivation is shown in Fig. 1 and Additional file 1 (Table S1). Non-response in the biomarker subsample was substantial: 43.8% of age-eligible children lacked a valid measurement, and these children differed systematically from those with a valid measurement (Additional file 1, Table S2). Rather than treat this as an unmodelled limitation, we specified the non-response mechanism explicitly and stress-tested the primary result under it (see Statistical analysis). Primary multivariable models used the 9,717 children with complete covariate information.

**Fig. 1.**
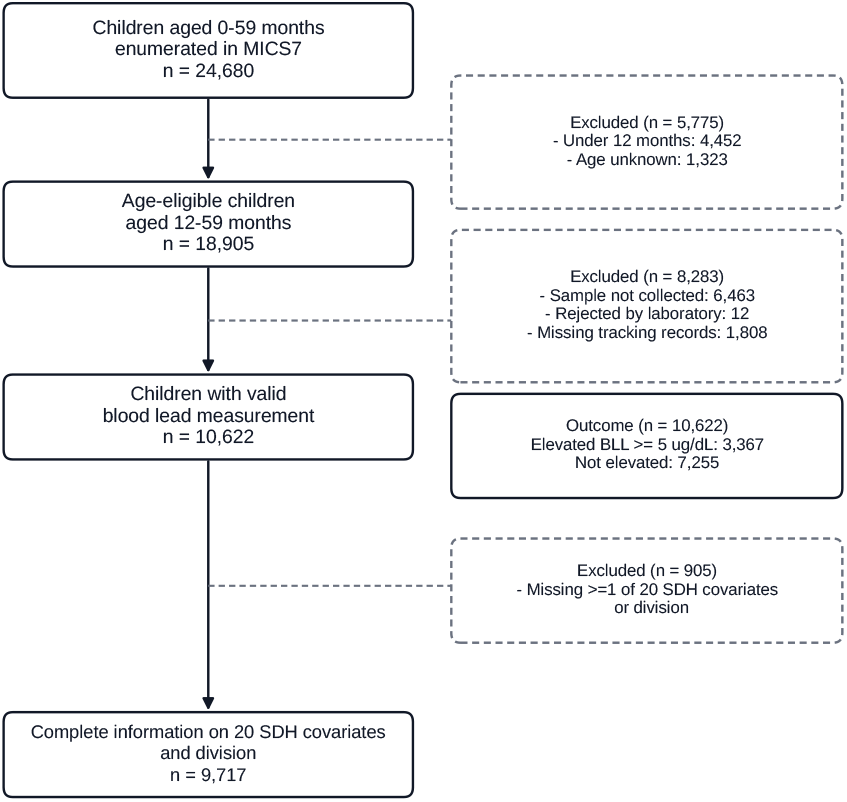
Participant flow. Of 24,680 children aged 0–59 months enumerated in MICS7 2025, 18,905 were age-eligible (12–59 months). A total of 8,283 lacked a valid blood lead measurement, yielding 10,622 children with biomonitoring data. Multivariable models were restricted to 9,717 children with complete covariate information (905 excluded for one or more missing SDH covariates or division).

### 2.3 Outcome and threshold justification

The primary outcome was BLL ≥ 5 µg/dL, retained for historical compatibility with prior Bangladeshi studies and with the UNICEF/Pure Earth threshold [15]. The current CDC blood lead reference value of ≥ 3.5 µg/dL was reported as a co-primary endpoint [12]. This value is derived from the upper tail (97.5th percentile) of the US child blood-lead distribution and functions as a screening and surveillance benchmark rather than a health-based threshold; no safe blood lead level in children has been identified. We use it here as a comparative benchmark for international context, not as a clinical or regulatory cutoff. Secondary endpoints were log-transformed continuous BLL and ≥ 10 µg/dL. The MICS7 documentation and laboratory partners do not report the assay method, laboratory quality-control statistics, or the limit of detection (LOD) for blood lead. The microdata contain five observations at the minimum reported value and a cluster of 67 children below 1.0 µg/dL, which is consistent with a LOD substitution rule (for example, 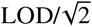 [32]), but the actual rule used for values below the LOD is not specified in the available documentation. We therefore used the concentrations as provided and cannot verify whether or how below-LOD values were substituted. This uncertainty may affect the continuous outcome and classification near the 3.5 µg/dL CDC reference value; we address this as a limitation.

### 2.4 Social determinants framework

Candidate predictors were structured following the WHO CSDH framework [33,34] and Victora’s hierarchical epidemiological framework [35]: (1) structural socioeconomic position (household wealth quintile, derived from the MICS asset-based wealth index [36, 37]; maternal education; household-head education); (2) social and cultural context (religion, ethnic minority status, women’s decision-making autonomy); geographic context (urban/rural residence, division); (4) parental and early-life factors (child age, sex, maternal age); (5) household material environment (crowding; improved floor, wall and sanitation); and (6) assets, healthcare and protection (refrigerator, bank account, agricultural land, livestock, dwelling ownership, safety-net transfers). Thirty candidate determinants were considered *a priori* from the WHO CSDH framework and Bangladesh-specific literature, subject to four pre-specified criteria: availability in the MICS7 microdata; missingness below 20% in the analytic sample; sufficient empirical variation; and absence of strong collinearity with an already-retained covariate. Twenty were retained; the ten exclusions and their reasons are listed in Additional file 1 (Table S17).

### 2.5 Statistical analysis

Our covariate set is a *descriptive predictor model*, not a causal identification strategy. Several covariates in the fully adjusted model — notably urban residence and household material environment — may lie on the pathway between wealth and BLL, and several asset and housing indicators are components of the MICS wealth index itself; adjusting for them yields *conditional associations* and, for index components, a suppression effect in which the wealth coefficient is estimated from residual variation [38]. The main tables report the crude and fully adjusted estimates; the complete staged sequence, including a model that excludes the dwelling indicator, is reported in Additional file 1 (Table S11).

Survey-weighted proportions and means with 95% confidence intervals (CIs) were estimated by Taylor series linearisation using the MICS7 stratum and PSU identifiers [39]; the same design-based machinery was used for every model, including the modified Poisson model, the inverse-probability weighting (IPW) analysis and each imputed dataset, so that all standard errors reflect the design consistently (design diagnostics: Additional file 1, Table S4). Multivariable survey logistic regression estimated adjusted odds ratios (aOR):

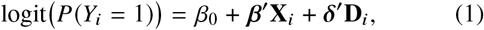

where **X**_*i*_ contains the 20 covariates and **D**_*i*_ denotes division fixed effects (reference: Barishal). Modified Poisson regression with robust sandwich standard errors estimated adjusted prevalence ratios (aPR) [40]. Because the outcome is common (38.5%), ORs can lie substantially further from the null than prevalence ratios [41], so we report staged models on both scales to assess suppression on a collapsible scale [42, 43]. Multicollinearity was assessed with variance inflation factors (VIF). Multiple-testing correction used the Benjamini–Hochberg procedure (*q* < 0.05) [44], applied separately to the aOR and aPR families. Per-1-SD odds ratios for continuous covariates are in Additional file 1 (Table S15). Departure of the wealth association from linearity was tested with a design-based Wald test (3 degrees of freedom), and heterogeneity of the wealth association across the eight divisions with a design-based Wald test (7 degrees of freedom) accompanied by division-stratified wealth aORs (Additional file 1, Table S12).

We assessed wealth × maternal-education interaction on the multiplicative scale (product term in the fully adjusted model, design-based Wald test) and on the additive scale (RERI, AP and S, with delta-method CIs [45, 46]). Because maternal education is preventive in this contrast, additive measures are reference-dependent [47]; we computed them under both reference codings and rely on the multiplicative interaction as the primary evidence. The full additive-scale reference-coding analysis is in Additional file 1 (Section S14, Table S14).

Sensitivity analyses addressed the non-response mechanism and robustness of the primary estimates. IPW used a survey-weighted logistic selection model fitted on the age-eligible frame [48] (Additional file 1, Table S7). Selection-bias scenarios multiplied the weight of elevated cases by a wealth-specific factor, including uniform factors (1.5, 2.0), adverse asymmetric scenarios (SB 2.0 in the poorest quintile only; SB 0.5 in the richest only), a symmetric worst case, and a tipping-point grid; these follow a pattern-mixture logic [49] (Additional file 1, Section S6, Table S6). Multiple imputation (*m* = 20; chained equations; Rubin/Barnard–Rubin pooling [50–52]) used the outcome, all 20 covariates, division and the log sampling weight (Table S8). 500 Rao–Wu bootstrap replicates [53] verified the Taylor standard errors (Table S5), and E-values on the prevalence-ratio scale quantified the unmeasured confounding needed to explain away the primary associations [54] (Table S9). Further specification checks (alternative outcomes, urban/rural strata, median imputation, unweighted analysis, exclusion of dwelling ownership) are in Table S10.

## 3 Results

### 3.1 Sample and overall prevalence

Of the 24,680 children enumerated in MICS7, 18,905 were age-eligible (12–59 months; Fig. 1). A valid blood lead measurement was available for 10,622 (56.2%), and complete covariate information for 9,717. In total, 3,367 children (31.7% unweighted) had elevated BLL, corresponding to a survey-weighted national prevalence of 38.5% (95% CI 37.2–39.9) at ≥5 µg/dL. At the CDC reference value of ≥ 3.5 µg/dL, 66.2% (64.9–67.4) of children exceeded the threshold. The survey-weighted median BLL was 4.32 µg/dL (weighted IQR 3.09–6.20; weighted mean 5.21 µg/dL); 7.1% of children had severe exposure (≥10 µg/dL). At both thresholds, a substantial share of children carry a body burden of lead at which neurodevelopmental effects have been documented in pooled analyses [4]. These estimates closely match those published in the final Survey Findings Report (Additional file 1, Table S3).

### 3.2 Main associations

Table 1 gives the unweighted descriptive picture; inference rests on the design-based models in Table 2. Elevated BLL was more frequent among children in the richest wealth quintile than the poorest (43.2% vs 27.1%), less frequent among children of mothers with tertiary than no formal education (25.3% vs 36.5%), and less frequent among owners of agricultural land, livestock and the dwelling (25.0%, 25.5% and 28.4%). Urban residence showed higher prevalence (40.9% vs 28.7% rural) but attenuated after full adjustment. Each of these associations was statistically significant (all *p* < 0.01).

**Table 1.** Distribution of social determinants by elevated blood lead status. (BLL ≥ 5 µg/dL), Bangladesh MICS7 2025 (*N* = 10,622)

| Variable | Category | Total |  | Elevated BLL |  | *p* |
| --- | --- | --- | --- | --- | --- | --- |
|  |  | *n* | % | *n* | row % |  |
| Structural socioeconomic position |  |  |  |  |  |  |
| Household wealth quintile | Q1 (poorest) | 2,823 | 26.6 | 766 | 27.1 | <0.001 |
|  | Q2 | 2,506 | 23.6 | 688 | 27.5 |  |
|  | Q3 | 2,144 | 20.2 | 663 | 30.9 |  |
|  | Q4 | 1,860 | 17.5 | 693 | 37.3 |  |
|  | Q5 (richest) | 1,289 | 12.1 | 557 | 43.2 |  |
| Maternal education | None | 750 | 7.1 | 274 | 36.5 | <0.001 |
|  | Primary | 2,049 | 19.3 | 766 | 37.4 |  |
|  | Lower sec. | 2,853 | 26.9 | 916 | 32.1 |  |
|  | Secondary | 2,926 | 27.5 | 875 | 29.9 |  |
|  | Higher sec. | 1,171 | 11.0 | 315 | 26.9 |  |
|  | Tertiary | 870 | 8.2 | 220 | 25.3 |  |
| Household-head education | Low | 5,961 | 56.1 | 1,925 | 32.3 | <0.001 |
|  | Mid | 3,173 | 29.9 | 1,032 | 32.5 |  |
|  | High | 1,481 | 13.9 | 407 | 27.5 |  |
| Social and cultural context |  |  |  |  |  |  |
| Religion | Muslim | 9,675 | 91.1 | 3,089 | 31.9 | 0.11 |
|  | Non-Muslim | 947 | 8.9 | 278 | 29.4 |  |
| Ethnicity | Bengali | 10,455 | 98.4 | 3,319 | 31.7 | 0.46 |
|  | Other | 167 | 1.6 | 48 | 28.7 |  |
| Women's autonomy | Low (≤0.67) | 2,807 | 26.4 | 896 | 31.9 | 0.73 |
|  | High (>0.67) | 7,518 | 70.8 | 2,371 | 31.5 |  |
| Geographic context |  |  |  |  |  |  |
| Residence | Rural | 7,982 | 75.1 | 2,287 | 28.7 | <0.001 |
|  | Urban | 2,640 | 24.9 | 1,080 | 40.9 |  |
| Division | Barishal | 1,088 | 10.2 | 292 | 26.8 | <0.001 |
|  | Chattogram | 2,120 | 20.0 | 833 | 39.3 |  |
|  | Dhaka | 1,790 | 16.9 | 1,029 | 57.5 |  |
|  | Khulna | 1,615 | 15.2 | 258 | 16.0 |  |
|  | Mymensingh | 658 | 6.2 | 171 | 26.0 |  |
|  | Rajshahi | 1,148 | 10.8 | 169 | 14.7 |  |
|  | Rangpur | 1,524 | 14.3 | 289 | 19.0 |  |
|  | Sylhet | 679 | 6.4 | 326 | 48.0 |  |

| Variable | Category | Total |  | Elevated BLL |  | *p* |
| --- | --- | --- | --- | --- | --- | --- |
|  |  | *n* | % | *n* | row % |  |
| Parental and early-life factors |  |  |  |  |  |  |
| Child sex | Female | 5,048 | 47.5 | 1,511 | 29.9 | <0.001 |
|  | Male | 5,574 | 52.5 | 1,856 | 33.3 |  |
| Child age (months) | Q1 [12–23] | 2,684 | 25.3 | 963 | 35.9 | <0.001 |
|  | Q2 [24–35] | 2,774 | 26.1 | 876 | 31.6 |  |
|  | Q3 [36–47] | 2,674 | 25.2 | 807 | 30.2 |  |
|  | Q4 [48–59] | 2,490 | 23.4 | 721 | 29.0 |  |
| Maternal age (years) | Q1 [15–22] | 2,879 | 27.1 | 930 | 32.3 | 0.90 |
|  | Q2 [23–26] | 2,498 | 23.5 | 789 | 31.6 |  |
|  | Q3 [27–31] | 2,515 | 23.7 | 792 | 31.5 |  |
|  | Q4 [32–49] | 2,575 | 24.2 | 812 | 31.5 |  |
| Household material environment |  |  |  |  |  |  |
| Crowding (per room) | Q1 [$\leq 2.0$] | 3,496 | 32.9 | 1,004 | 28.7 | <0.001 |
|  | Q2 [2.1–2.5] | 2,381 | 22.4 | 721 | 30.3 |  |
|  | Q3 [2.6–3.3] | 2,398 | 22.6 | 789 | 32.9 |  |
| | Q4 [$> 3.3$] | 2,347 | 22.1 | 853 | 36.3 | |
| Improved floor | No | 5,628 | 53.0 | 1,572 | 27.9 | <0.001 |
|  | Yes | 4,994 | 47.0 | 1,795 | 35.9 |  |
| Improved wall | No | 6,117 | 57.6 | 1,928 | 31.5 | 0.66 |
|  | Yes | 4,505 | 42.4 | 1,439 | 31.9 |  |
| Improved sanitation | No | 1,156 | 10.9 | 426 | 36.9 | <0.001 |
|  | Yes | 9,466 | 89.1 | 2,941 | 31.1 |  |
| Assets, healthcare and protection |  |  |  |  |  |  |
| Refrigerator | No | 4,538 | 42.7 | 1,374 | 30.3 | 0.003 |
|  | Yes | 5,798 | 54.6 | 1,915 | 33.0 |  |
| Bank account | No | 5,724 | 53.9 | 1,823 | 31.8 | 0.74 |
|  | Yes | 4,898 | 46.1 | 1,544 | 31.5 |  |
| Agricultural land | No | 6,933 | 65.3 | 2,444 | 35.3 | <0.001 |
|  | Yes | 3,686 | 34.7 | 923 | 25.0 |  |
| Livestock | No | 3,804 | 35.8 | 1,630 | 42.8 | <0.001 |
|  | Yes | 6,818 | 64.2 | 1,737 | 25.5 |  |
| Dwelling | No | 930 | 8.8 | 588 | 63.2 | <0.001 |
|  | Yes | 9,474 | 89.2 | 2,694 | 28.4 |  |
| Safety-net transfer | No | 6,539 | 61.6 | 2,233 | 34.1 | <0.001 |
|  | Yes | 3,948 | 37.2 | 1,098 | 27.8 |  |

**Table 2.** Multivariable survey-weighted associations between social determinants and elevated childhood blood lead. (BLL≥5 µg/dL), Bangladesh MICS7 2025

| Domain and covariate | n | Prev. % | Odds ratio (95% CI) |  | Prevalence ratio | q |
| --- | --- | --- | --- | --- | --- | --- |
|  |  |  | COR | aOR<sub>2</sub> | aPR (95% CI) |  |
| Domain 1 - Structural socioeconomic position |  |  |  |  |  |  |
| Household wealth (per quintile) | 10,622 | 49.4 | 1.29 (1.24–1.34) | 1.24 (1.13–1.37)*** | 1.11 (1.06–1.17)*** | <0.001 |
| Maternal education (per level) | 10,619 | 34.8 | 0.88 (0.85–0.91) | 0.83 (0.79–0.88)*** | 0.92 (0.89–0.94)*** | <0.001 |
| Household-head education (per level) | 10,615 | 37.1 | 0.99 (0.96–1.02) | 1.00 (0.95–1.04) | 1.00 (0.98–1.03) | 0.966 |
| Domain 2 - Social and cultural context |  |  |  |  |  |  |
| Non-Muslim household (ref. Muslim) | 10,622 | 35.4 | 0.87 (0.68–1.10) | 0.91 (0.71–1.17) | 0.95 (0.83–1.09) | 0.621 |
| Ethnic minority (ref. Bengali) | 10,622 | 26.6 | 0.58 (0.36–0.92)* | 1.03 (0.48–2.20) | 1.02 (0.62–1.67) | 0.966 |
| Women's autonomy index (0–1) | 10,325 | 38.0 | 0.88 (0.76–1.02) | 1.00 (0.85–1.17) | 1.02 (0.94–1.10) | 0.966 |
| Domain 3 - Geographic context |  |  |  |  |  |  |
| Urban residence (ref. rural) | 10,622 | 49.9 | 1.95 (1.72–2.22)*** | 1.07 (0.91–1.26) | 1.04 (0.96–1.12) | 0.621 |
| Domain 4 - Parental and early-life factors |  |  |  |  |  |  |
| Child age (per month) | 10,622 | 35.9 | 0.994 (0.990–0.998)** | 0.993 (0.988–0.997)** | 0.996 (0.994–0.999)** | 0.004 |
| Male child (ref. female) | 10,622 | 40.5 | 1.19 (1.08–1.31)*** | 1.28 (1.14–1.44)*** | 1.13 (1.07–1.20)*** | <0.001 |
| Maternal age (per year) | 10,467 | 37.1 | 0.992 (0.985–1.000) | 0.986 (0.976–0.995)** | 0.993 (0.989–0.998)** | 0.007 |
| Domain 5 - Household material environment |  |  |  |  |  |  |
| Crowding (persons per room) | 10,622 | 42.9 | 1.19 (1.13–1.26)*** | 1.08 (1.01–1.15)* | 1.03 (1.00–1.06) | 0.049 |
| Improved floor material | 10,622 | 45.1 | 1.82 (1.63–2.02)*** | 0.98 (0.82–1.17) | 0.99 (0.89–1.09) | 0.966 |
| Improved wall material | 10,622 | 41.7 | 1.28 (1.15–1.42)*** | 0.82 (0.69–0.96)* | 0.89 (0.82–0.97)* | 0.038 |
| Improved sanitation facility | 10,622 | 37.5 | 0.69 (0.59–0.82)*** | 0.97 (0.82–1.16) | 0.99 (0.91–1.07) | 0.961 |
| Domain 6 - Assets, healthcare and protection |  |  |  |  |  |  |
| Refrigerator ownership | 10,336 | 39.9 | 1.14 (1.03–1.27)* | 0.90 (0.77–1.04) | 0.95 (0.89–1.02) | 0.232 |
| Bank account ownership | 10,622 | 39.1 | 1.05 (0.94–1.16) | 0.91 (0.79–1.04) | 0.96 (0.90–1.02) | 0.244 |
| Agricultural land ownership | 10,619 | 29.0 | 0.54 (0.49–0.61)*** | 0.77 (0.67–0.88)*** | 0.86 (0.79–0.93)*** | <0.001 |
| Livestock ownership | 10,622 | 29.1 | 0.39 (0.35–0.43)*** | 0.77 (0.67–0.88)*** | 0.86 (0.80–0.93)*** | <0.001 |
| Dwelling ownership | 10,404 | 33.0 | 0.20 (0.17–0.24)*** | 0.54 (0.43–0.69)*** | 0.84 (0.77–0.93)*** | <0.001 |
| Social safety-net transfer | 10,487 | 32.2 | 0.66 (0.59–0.73)*** | 0.90 (0.79–1.02) | 0.94 (0.88–1.01) | 0.158 |

Note: COR, crude odds ratio; aOR<sub>2</sub>, fully adjusted for all 20 covariates plus division ( $n = 9,717$ ); aPR, adjusted prevalence ratio from a modified Poisson model with survey-weighted sandwich variance; q, Benjamini–Hochberg false discovery rate for the aOR<sub>2</sub> model. Bold denotes FDR-significant covariates in the fully adjusted model. The full staged model sequence is reported in Additional file 1 (Table S11). Prev. %, weighted prevalence among the exposed group (binary: yes; continuous: top quartile); shown for reference only. Per-1-SD odds ratios for continuous covariates are in Additional file 1 (Table S15). Significance: \* $q < 0.05$ ; \*\* $q < 0.01$ ; \*\*\* $q < 0.001$ .

In the fully adjusted model (Table 2; Fig. 2), ten of 20 covariates survived Benjamini–Hochberg correction. Maternal education was the most precisely estimated inverse association (aOR_2_ 0.83 per level, 95% CI 0.79–0.88; aPR 0.92, 0.89– 0.94; *q* < 0.001), followed by improved wall construction (0.82, 0.69–0.96; *q* = 0.038). Crowding (1.08, 1.01–1.15; *q* = 0.049) and male sex (1.28, 1.14–1.44; *q* < 0.001) were positively associated with elevated BLL. Age was inversely associated on both axes: each additional month of child age reduced the odds of elevated BLL by approximately 0.7% (aOR_2_ 0.993, 0.988–0.997; *q* = 0.004), and each additional year of maternal age reduced the odds by approximately 1.4% (0.986, 0.976–0.995; *q* = 0.007). Both per-unit effects are small, but the implied contrasts across the observed age range are non-trivial: a child aged 59 months versus 12 months had approximately 28% lower adjusted odds of elevated BLL, and a mother aged 45 years versus 18 years was associated with approximately 32% lower adjusted odds in her child. Three agrarian assets showed independent inverse associations: agricultural land (0.77, 0.67–0.88), livestock (0.77, 0.67–0.88) and the dwelling itself (0.54, 0.43–0.69; all *q* < 0.001); the dwelling estimate is furthest from the null but is a binary ownership contrast and is not directly comparable to the per-level maternal-education coefficient. Improved flooring and sanitation were null in the fully adjusted model, and refrigerator, bank-account and safety-net variables were fully attenuated once household wealth entered the model (weighted prevalences by covariate: Additional file 1, Table S16). Cultural factors (religion, ethnicity, women’s autonomy) showed no independent association (all *q* > 0.6). Urban residence was strongly elevated univariately (COR 1.95, 1.72–2.22) but fully attenuated after adjustment (aOR_2_ 1.07, 0.91–1.26; *q* = 0.621), consistent with the causal-framing caveat in the Methods.

**Fig. 2.**
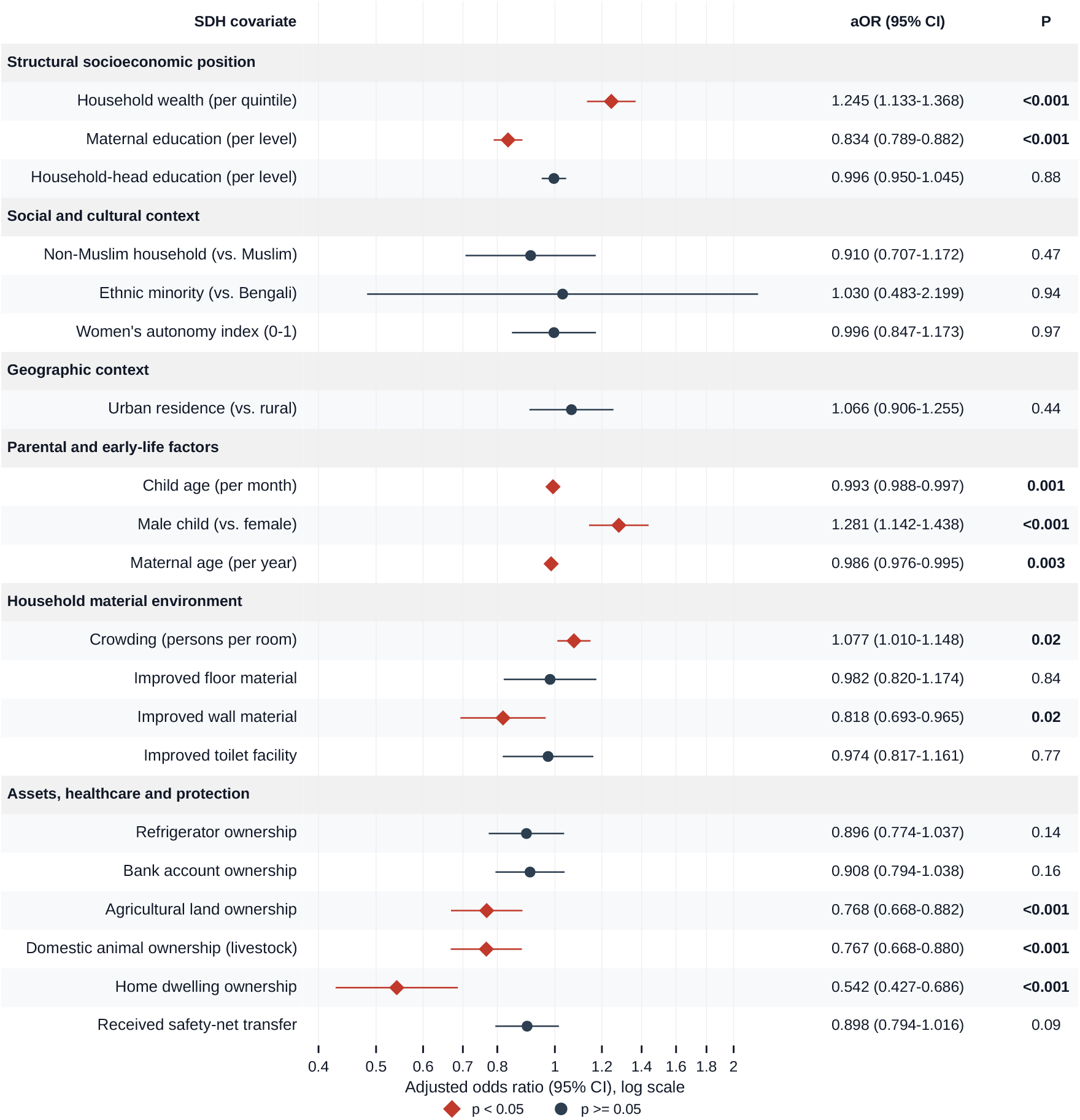
Adjusted odds ratios for social determinants of elevated blood lead by WHO CSDH domain. (*n* = 9,717). Point estimates (aOR_2_) and 95% CIs from the fully adjusted model with division fixed effects. Wealth points to the right (harmful); maternal education and agrarian capital point to the left (inverse), jointly reflecting the reverse socioeconomic gradient observed in this setting.

### 3.3 The reverse wealth gradient and its sub-national heterogeneity

#### Gradient

Prevalence rose monotonically with household wealth, from 30.2% (95% CI 28.0–32.6) in the poorest quintile to 51.9% (48.8–55.0) in the richest, with intermediate values of 30.5% (Q2), 36.7% (Q3) and 47.3% (Q4); the survey-weighted median BLL rose in parallel from 3.97 to 5.15 µg/dL (Fig. 3). Each quintile increase in wealth was associated with a fully adjusted aOR_2_ 1.24 (1.13–1.37; aPR 1.11, 1.06–1.17; *q* < 0.001). The crude association is already positive (1.29, 1.24–1.34; Table 2), so no adjustment set reverses its sign, and the fully adjusted estimate is similar in magnitude to the crude. The complete staged sequence — from a minimal age/sex/division adjustment through each successive covariate block — is reported in Additional file 1 (Table S11).

**Fig. 3.**
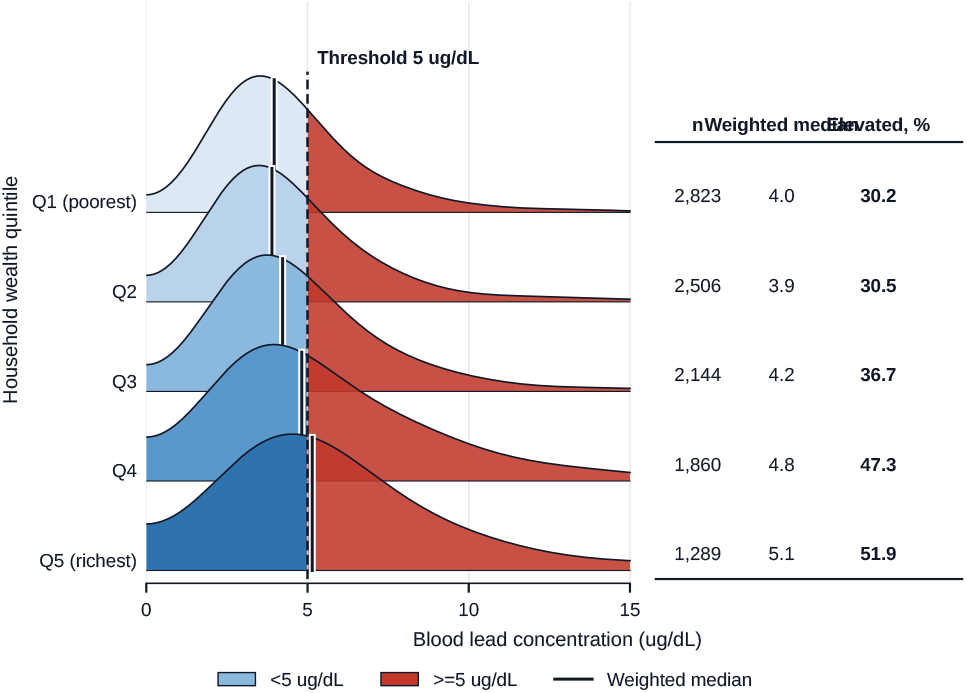
Survey-weighted blood lead density by wealth quintile. (*N* = 10,622). The dashed line marks the 5 µg/dL threshold. Distributions shift progressively rightward from the poorest (Q1) to the richest (Q5) quintile.

#### Geographic variation and heterogeneity

Division-level prevalence varied widely, from 16.0% (13.3–19.0) in Rajshahi to 65.6% (62.8–68.2) in Dhaka (Table 3; district-level variation in Additional file 1, Fig. S1; division forest plot, Additional file 1, Fig. S2). These figures agree closely with the corresponding division-level prevalences published in the final Survey Findings Report (29.0%, 42.1%, 65.2%, 17.8%, 30.6%, 15.9%, 19.7%, 46.8%, for Barishal, Chattogram, Dhaka, Khulna, Mymensingh, Rajshahi, Rangpur and Sylhet respectively; Additional file 1, Table S3). Relative to Barishal (29.0%, 25.3–33.1), children in Dhaka had approximately three-fold adjusted odds of elevated lead (aOR_2_ 2.99, 2.34–3.82; aPR 1.76, 1.52–2.05), and Sylhet and Chattogram were also elevated, whereas Khulna, Rangpur and Rajshahi had significantly reduced odds; Mymensingh was indistinguishable from Barishal. The Dhaka excess persisted after adjustment for wealth, urban residence and crowding; we did not measure the geographic distribution of lead sources, so any explanation (for example, a greater concentration of consumer infrastructure in the capital) is hypothesis-generating. Critically, the wealth gradient itself was significantly heterogeneous across divisions (Wald 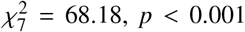): strongest in Dhaka (aOR 1.70, 1.52–1.90), smaller but with a confidence interval excluding 1 in Khulna (1.20, 1.05–1.38) and Barishal (1.16, 1.01–1.34), positive but imprecise in Mymensingh (1.23, 0.97–1.57), and null in Chattogram, Rajshahi, Rangpur and Sylhet (Table 3; Additional file 1, Table S12).

**Table 3.** Division-specific prevalence-ratio and odds-ratio associations with elevated childhood blood lead (relative to Barishal) and division-stratified wealth gradients.

| Division | n valid | n cc | Division vs. Barishal |  | Wealth gradient within division |
| --- | --- | --- | --- | --- | --- |
|  |  |  | aOR<sub>2</sub> (95% CI) | aPR (95% CI) | Wealth aOR per quintile (95% CI) |
| Barishal (ref.) | 1,088 | 1,001 | 1.00 (ref.) | 1.00 (ref.) | 1.16 (1.01–1.34) |
| Chattogram | 2,120 | 1,812 | 1.34 (1.04–1.73)* | 1.25 (1.07–1.47) | 1.04 (0.93–1.16) |
| Dhaka | 1,790 | 1,689 | **2.99 (2.34–3.82)\*\*\*** | **1.76 (1.52–2.05)** | **1.70 (1.52–1.90)** |
| Khulna | 1,615 | 1,538 | 0.53 (0.41–0.69)*** | 0.63 (0.52–0.76) | 1.20 (1.05–1.38) |
| Mymensingh | 658 | 619 | 0.96 (0.70–1.32) | 1.00 (0.81–1.24) | 1.23 (0.97–1.57) |
| Rajshahi | 1,148 | 1,032 | 0.42 (0.31–0.57)*** | 0.53 (0.42–0.66) | 0.97 (0.79–1.17) |
| Rangpur | 1,524 | 1,398 | 0.61 (0.47–0.79)*** | 0.69 (0.57–0.83) | 1.03 (0.91–1.18) |
| Sylhet | 679 | 628 | 1.76 (1.31–2.36)*** | 1.45 (1.22–1.73) | 0.96 (0.81–1.16) |

Note: aOR<sub>2</sub> and aPR adjust for all 20 covariates. $n$ valid = all children with valid BLL in that division (sums to 10,622); $n$ cc = complete-case subsample contributing to the model (sums to 9,717). Wealth aORs are division-stratified using a reduced adjustment set (maternal education, urban residence, crowding, child age, child sex) to preserve stability in the smaller division subsamples; the pooled wealth coefficient in Table 2 is fully adjusted for all 20 SDH covariates. The formal test of heterogeneity across divisions was Wald $\chi^2 = 68.18$ , $p < 0.001$ . Significance (division contrasts): \* $p < 0.05$ ; \*\* $p < 0.01$ ; \*\*\* $p < 0.001$ .

#### Wealth × maternal-education interaction

Prevalence rose with wealth inside every education stratum and fell with education inside every wealth quintile (Fig. 4). The wealth association diminished with maternal education: the rise from the lowest to the highest wealth quintile with *n* ≥50 was +43.6 percentage points (pp) among children of mothers with no formal education (Q1 to Q4) and +43.9 pp among those with primary education, falling to +17.1 pp among tertiary-educated mothers (Q2 to Q5); values for the intermediate strata are in Additional file 1 (Table S13). Formally, the wealth × maternal-education interaction was significant on the multiplicative scale (product-term aOR 0.945, 95% CI 0.913–0.977; design-based Wald *p* = 0.001). Additive-scale measures were reference-dependent and yielded opposite signs under the two possible reference codings; we therefore base no substantive claim on the additive scale (Additional file 1, Table S14). Within the higher-secondary stratum the per-quintile wealth aPR was 1.09 (1.02–1.16), and within the tertiary stratum it was 1.07 (1.00–1.15); the education association among rich households was 0.82 (0.75–0.88).

**Fig. 4.**
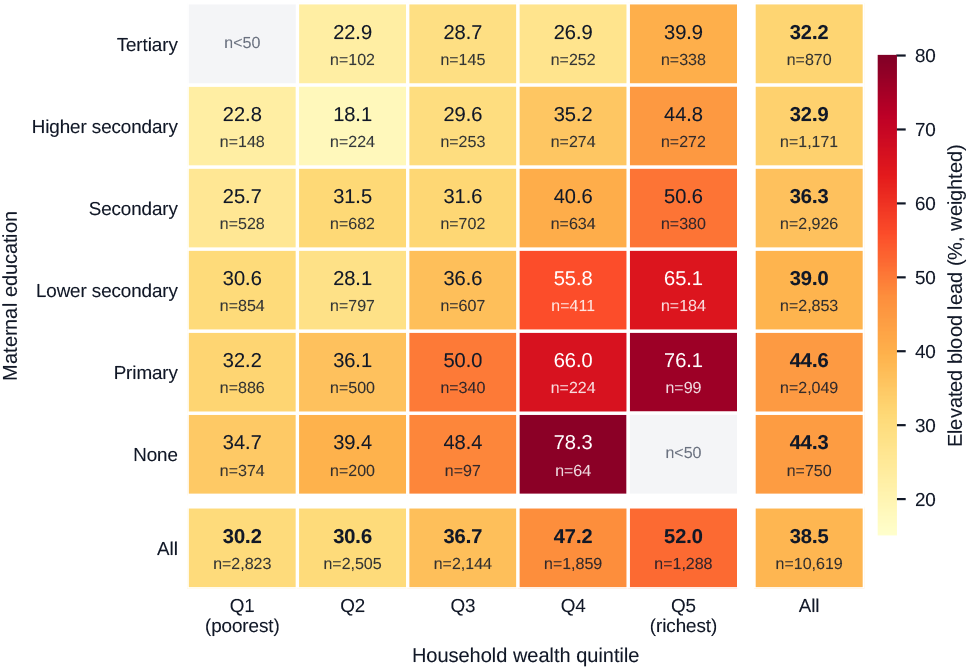
Elevated-BLL prevalence (%) by wealth quintile and maternal education. Denominators are the 10,619 children with non-missing wealth and maternal education (3 fewer than the full valid-BLL sample of Table 1). Prevalence rises with wealth and declines with maternal education. Cells with *n* < 50 are suppressed (grey). The wealth association attenuates but is not eliminated at higher maternal education.

Under the alternative education cut-point, the wealth aPR within the high-education stratum was 1.08 (0.94–1.25), with the confidence interval including 1 (Additional file 1, Table S14). We therefore describe maternal education as *attenuating but not eliminating* the wealth association, and we do not claim full neutralisation. The analysis measures neither household behaviour nor market-mediated exposure, so the residual association at higher education levels is an association, not evidence about any specific mechanism.

### 3.4 Sensitivity analyses

The primary result was stress-tested under the non-response mechanism specified in the Methods (Additional file 1, Tables S5–S10; Fig. 5). IPW returned a wealth aOR of 1.260 (Taylor 1.139–1.394) and an IPW-adjusted prevalence of 41.5% (40.1–43.0) — about 3 percentage points *above* the headline 38.5% — indicating that, under the specified selection model, the naive estimate is biased downwards and the primary findings are conservative with respect to the wealth gradient; multiple imputation gave a pooled wealth aOR of 1.247 (1.139–1.365). Under uniform selection-bias factors of 1.5 and 2.0 the wealth aOR moved only from 1.245 to 1.238 and 1.233. The association did not remain positive and significant under the two adverse asymmetric scenarios (SB 2.0 in the poorest quintile only: aOR 1.041, 0.948–1.144; SB 0.5 in the richest only: 1.098, 0.998–1.207), and the point estimate crossed the null under the symmetric worst case (aOR 0.915, 0.833–1.006); the tipping-point grid first crossed aOR ≤ 1 at *r* = 1.75. Bootstrap standard errors matched Taylor standard errors to within 6.9%, and E-values indicated that an unmeasured confounder would need prevalence-ratio associations of 1.47 (CI limit 1.31) with both wealth and the outcome to explain away the wealth gradient. Estimates were stable across alternative outcomes, urban and rural strata, missing-data handling, weighting and exclusion of dwelling ownership (Table S10); the maximum VIF was 4.11 (Table S4).

**Fig. 5.**
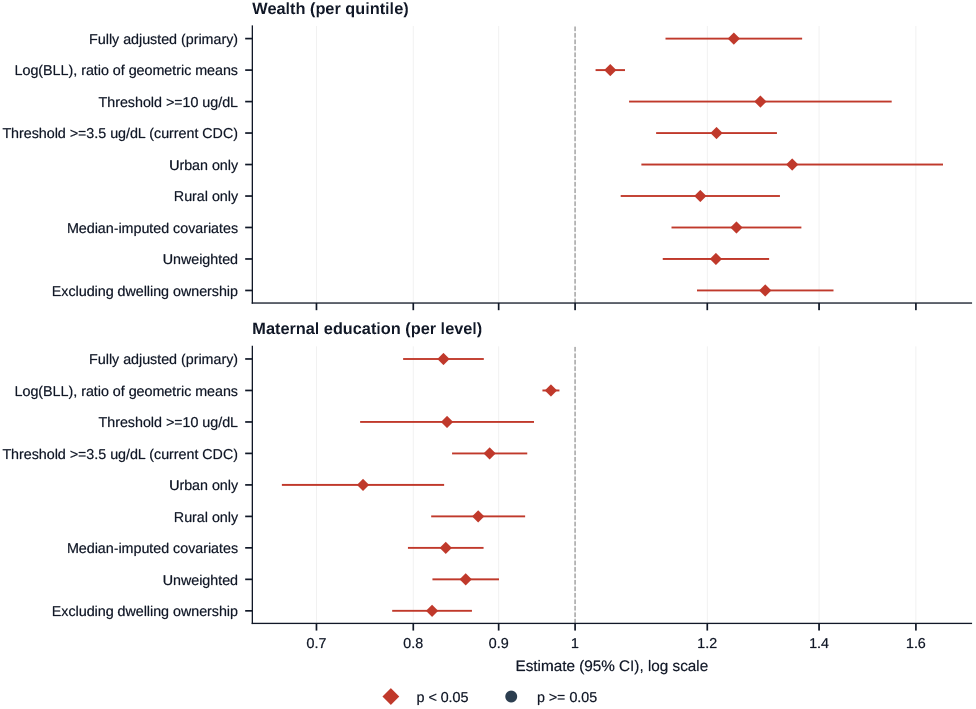
Sensitivity forest plot. Wealth aOR (per quintile) across the primary model, alternative outcomes, subpopulation strata, missing-data handling, weighting and design specifications, and selection-bias scenarios. The dashed vertical line marks the primary estimate (aOR 1.245). The wealth association remains positive across standard and favourable scenarios; it attenuates under the two adverse asymmetric selection-bias scenarios and its point estimate crosses the null under the symmetric worst case.

## 4 Discussion

### 4.1 Principal findings

This is the first nationally representative picture of paediatric blood lead in Bangladesh, and the story it tells is the opposite of the classic environmental-justice one. Children in wealthier households are more exposed, not less. Elevated blood lead affected 38.5% of children aged 12–59 months at the ≥5 µg/dL threshold, and 66.2% at the CDC blood lead reference value of ≥3.5 µg/dL — a screening and surveillance benchmark derived from the upper tail of the US child blood-lead distribution, not a health-based threshold. Prevalence rose monotonically with household wealth, from 30.2% in the poorest quintile to 51.9% in the richest. The crude wealth association is already positive (aOR 1.29, 1.24–1.34), so no adjustment set reverses its sign; the fully adjusted estimate is similar in magnitude (1.24, 1.13–1.37). The complete staged sequence is reported in Additional file 1 (Table S11). Several classic determinants behaved as expected: maternal education was strongly protective (aOR 0.83 per level), improved wall construction reduced risk, household crowding increased it, boys were more exposed than girls (aOR 1.28), and older children and children of older mothers had modestly lower odds (per-month aOR 0.993; per-year 0.986). The age and sex gradients are consistent with the paediatric lead literature, in which younger children absorb a larger fraction of an ingested dose and boys have higher hand-to-mouth activity. Agrarian capital (ownership of agricultural land, livestock and the dwelling) formed the most robustly protective set of assets. Formal testing of the wealth × education interaction was significant on the multiplicative scale (product-term aOR 0.945, 0.913– 0.977; design-based Wald *p* = 0.001) and on the additive scale under the alternative education cut-point (RERI −0.18, *p* = 0.02); under the main cut-point the additive interaction was borderline (RERI−0.14, *p* = 0.06). The wealth effect within the higher-secondary stratum remained positive (per-quintile aPR 1.09, 1.02–1.16), and the tertiary-stratum estimate was similar but with a confidence interval including 1 (1.07, 1.00–1.15), indicating that maternal education attenuates but does not eliminate the wealth penalty. The reverse gradient was resilient across IPW, multiple imputation, six non-reference selection-bias scenarios and alternative outcome definitions; the point estimate crossed 1.0 only when elevated-case weights in the poorest quintile were inflated 1.75-fold relative to the richest. Sub-nationally, Dhaka Division showed approximately three-fold adjusted odds relative to Barishal.

### 4.2 Comparison with previous studies

Earlier investigations of Bangladeshi children relied on convenience samples. School-based assessments in Dhaka reported mean BLLs of 15.0 µg/dL [26], and sampling in the Tongi industrial belt documented near-universal levels above 10 µg/dL [27]. These studies identified hotspots but could not describe national patterning. Our weighted national median of 4.32 µg/dL is lower than the Dhaka school-based mean; however, a weighted national median and a single-city convenience-sample mean are not directly comparable, and we therefore make no claim about temporal decline. Our findings are compatible with the plausibility of food-chain and consumer-product pathways documented in South Asia: turmeric adulteration with lead chromate has been demonstrated in Bangladesh [20, 22], and commercial spices have been identified as a measurable source of childhood exposure [21]; however, those studies identify sources, not social gradients. A study of 135 children from households proximal or distal to battery-recycling operations in Patna, Bihar, found that 87% had BLLs above 5 µg/dL [55]; its sampling frame was not designed to be representative, so the estimate is not a general-population prevalence. Where prior sub-national studies in Bangladesh could identify hotspots but not national determinants, our analysis provides the first nationally representative quantification of the social patterning of childhood blood lead. Three elements are novel: a formal test of divisional heterogeneity of the wealth–BLL association using nationally representative data, an explicit multiplicative and additive-scale evaluation of wealth × maternal-education interaction with reference-coding transparency, and the whole quantification structured within the WHO CSDH framework so that results are directly comparable across settings.

### 4.3 Possible mechanisms and policy implications

The MICS7 data measure blood lead and social determinants but not dietary intake, product purchases or source-specific exposure; consequently, every mechanism discussed here is a hypothesis supported by regional literature, not a finding of these data. Informal used lead-acid battery recycling is among the most frequently reported probable sources of childhood lead exposure in LMICs [13], and the Savar study identifies local industrial exposure as a plausible pathway [24]; however, it describes a community-level exposure around a specific site rather than a socioeconomic pattern. Market-mediated sources are also documented: turmeric adulterated with lead chromate is well documented in Bangladesh [20, 21], although a 2017–2021 multi-component intervention and enforcement effort was followed by a decline in detectable lead at the largest wholesale market from 47% (2019) to 0% (2021) [57]. Lead-containing foodware and paint have also been identified in LMIC market screenings [23]. Whether higher-income Bangladeshi households purchase contaminated goods in greater volume or at higher unit value is not established and remains an untested hypothesis for source-apportionment work. The inverse associations with land and livestock ownership do not establish that subsistence production reduces exposure: work among pregnant women in rural Bangladesh has implicated several competing pathways, including lead-soldered cans, certain pesticide brands and rice grinding [58], and differences in the composition of purchased goods could equally explain the pattern. The persistence of a positive wealth association at higher maternal education is consistent with education operating through procurement discernment and health literacy, but also with diminishing returns to wealth at the top of the education distribution. These findings do not imply that wealth causes lead exposure or that protection should be redirected away from the poorest; they imply that exposure is not confined to the poorest households, so surveillance and product regulation should reach the broader population in parallel with continued protection of the poorest.

### 4.4 Strengths and limitations

Strengths of this study include the nationally representative sampling frame, an *a priori* CSDH framework, design-based inference with modified Poisson estimation of prevalence ratios, the formal test of divisional heterogeneity of the wealth– BLL association that earlier sub-national studies could not perform [26, 27], and a comprehensive set of sensitivity analyses (Additional file 1). Limitations warrant acknowledgment. **Design and confounding:** the cross-sectional design precludes causal attribution; several covariates (urban residence, housing materials, assets) are plausibly on the wealth–BLL pathway or are components of the wealth index, so the fully adjusted wealth coefficient is a conditional association, and the staged sequence (Table S11) is the appropriate reference for how the gradient evolves; the interaction describes joint patterning, not biological synergy, and additive-scale measures are reference-dependent when one exposure is preventive [47]. **Selection and missing data:** 43.8% of age-eligible children lacked a valid measurement and were systematically younger, wealthier (mean quintile 3.1 vs 2.7), more urban (29.6% vs 24.9%) and more concentrated in Dhaka and Chattogram (Table S2). The IPW-adjusted prevalence of 41.5% (40.1–43.0) is approximately 3 percentage points above the headline 38.5% (37.2–39.9), which under the specified selection model is consistent with the headline estimate being biased *downwards*; the non-overlapping intervals alone do not establish bias, so we present this as a model-based sensitivity estimate, not a demonstrated correction. The wealth gradient attenuated under the two adverse asymmetric scenarios and crossed the null under the symmetric worst case, and is therefore robust to most but not all tested specifications (Tables S6, S7, S10). **Exposure measurement:** MICS7 provides no isotope signatures, so individual-level source attribution is impossible; cooking-fuel data were unavailable in the child file, precluding analysis of indoor biomass combustion; blood lead came from a single laboratory assay; the assay method, laboratory quality-control statistics, and limit of detection were not reported in the available MICS7 documentation, and the rule used for values below the limit of detection is unknown. A cluster of 67 children below 1.0 µg/dL is consistent with a substitution rule, but we could not verify how those values were generated. This uncertainty may affect the continuous outcome and classification near the 3.5 µg/dL CDC reference value. If misclassification of the binary outcome is non-differential with respect to the exposures, error near the 5 µg/dL threshold would tend to bias estimates towards the null; other patterns could bias in either direction. **Generalisability:** the MICS wealth index is a relative asset measure whose quintile cut-points are not comparable across countries, and DHS/MICS asset indices have a documented urban bias [59, 60]; as urban and consumer-market pathways are central to our interpretation, we describe the result as a *relative-asset or consumer-position* gradient rather than a pure wealth gradient. The wealth association was modelled as monotonic across quintiles; tests of non-linearity did not reject linearity (*p* = 0.29 and *p* = 0.65), but the study was not powered to detect small departures from linearity.

## 5 Conclusion

This national analysis shows that elevated blood lead among Bangladeshi children aged 12–59 months follows a reverse socioeconomic gradient: wealthier children were more exposed, while maternal education and agrarian assets were inversely associated. The wealth association varied significantly across divisions (Wald 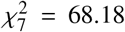, *p* < 0.001), with the largest excess in Dhaka, a formal test that earlier sub-national studies could not perform. Maternal education attenuated but did not eliminate the wealth association (multiplicative product-term aOR 0.945; per-quintile aPR 1.09 within the higher-secondary stratum). The wealth association was robust to most, but not all, tested selection-bias specifications: it attenuated under adverse asymmetric scenarios and its point estimate crossed the null under the symmetric worst case. These findings describe the social pattern of exposure, not its source; market-mediated and industrial pathways remain hypotheses that will require isotope studies and household-level exposure assessment, which MICS7 does not provide. Lead surveillance and risk communication should therefore extend beyond poverty-targeted frameworks to middle- and high-income communities, alongside continued protection of the poorest.

## Data Availability

The datasets analysed during the current study are available in the UNICEF Multiple Indicator Cluster Surveys repository under a data access agreement, following registration and approval of a data access request. The complete Python code for data preparation, statistical models, and figure generation is provided as a supplementary file.

https://mics.unicef.org/surveys

## Abbreviations

aOR: adjusted odds ratio
AP: attributable proportion
aPR: adjusted prevalence ratio
BBS: Bangladesh Bureau of Statistics
BLL: blood lead level
CDC: Centers for Disease Control and Prevention
CI: confidence interval
COR: crude odds ratio
CSDH: Commission on Social Determinants of Health
FDR: false discovery rate
IPW: inverse-probability-of-selection weighting
LMIC: low- and middle-income country
MI: multiple imputation
MICS: Multiple Indicator Cluster Survey
MNAR: missing not at random
pp: percentage points
PSU: primary sampling unit
RERI: relative excess risk due to interaction
S: synergy index
SB: selection-bias factor
SD: standard deviation
SDH: social determinants of health
STROBE: Strengthening the Reporting of Observational Studies in Epidemiology
UNICEF: United Nations Children’s Fund
VIF: variance inflation factor
WHO: World Health Organization

## Declarations

## Acknowledgements

The author thanks UNICEF Bangladesh and the Bangladesh Bureau of Statistics for conducting the survey and facilitating data access.

## Authors’ contributions

S.T.H. conceptualised the study, prepared and analysed the dataset, generated all models and figures, and drafted the manuscript. The author approved the final version.

## Data availability

The datasets analysed are available in the UNICEF MICS repository (https://mics.unicef.org/surveys) [61]. Analysis scripts supporting the findings are provided as Additional file 2. The STROBE checklist completed for this study is provided as Additional file 3.

## Ethics approval and consent to participate

The MICS7 survey protocol was approved by the Technical and Ethical Review Committee of the Bangladesh Bureau of Statistics and by the icddr,b Institutional Review Board (Protocol Reference PR-24113). All participants provided informed consent prior to data collection. This secondary analysis of de-identified data was exempt from additional institutional ethics review under University of Dhaka policy for research on non-identifiable secondary data.

## Consent for publication

Not applicable.

## Use of AI

During the preparation of this work, the author used a large language model to assist with language editing, LaTeX formatting, and manuscript organisation. The author reviewed and edited all AI-assisted content, takes full responsibility for the content of the publication, and confirms that no AI tool was used to generate research data, analyse data, or produce scientific conclusions.

## Author details

^1^Institute of Health Economics, University of Dhaka, Dhaka 1000, Bangladesh.

## Additional files

**Additional file 1** (PDF). Supplementary Tables S1–S17 and Figures S1–S2: analytic-sample derivation (S1); missing counts and denominators (S2); concordance with the final Survey Findings Report (S3); design diagnostics — ICC, design effect and VIF (S4); Rao–Wu bootstrap validation (S5); selection-bias scenarios with tipping-point grid (S6); IPW construction and estimates (S7); multiple-imputation diagnostics (S8); E-values (S9); robustness of the wealth and maternal-education associations across alternative specifications (S10); staged wealth models (S11); division-stratified wealth gradients (S12); wealth × maternal-education cell prevalences (S13); additive-scale interaction under both reference codings and stratum-specific estimates (S14); per-SD odds ratios (S15); weighted prevalences and design-based Wald tests by covariate category (S16); candidate-variable screening (S17); district choropleth (Fig. S1); forest plot of division odds ratios (Fig. S2).

**Additional file 2** (ZIP). Complete Python analysis and replication pipeline (<monospace>replication_script.py</monospace>) reproducing every table, figure and sensitivity analysis in this manuscript.

**Additional file 3** (PDF). Completed STROBE checklist for cross-sectional studies.

